# An adapted health system access framework for avoidable inflammatory bowel disease admissions: A scoping review

**DOI:** 10.1101/2025.09.08.25335323

**Authors:** Rachel L Hawkins, Kimberly Butler, Fiona Sampson, Alan J Lobo, Daniel Hind

**Affiliations:** Sheffield Centre for Health and Related Research, School of Medicine and Population Health, University of Sheffield, Sheffield, United Kingdom; Sheffield Inflammatory Bowel Disease Centre, Sheffield Teaching Hospitals NHS Foundation Trust, Sheffield, Sheffield, United Kingdom; School of Healthcare, The University of Leeds, Leeds, United Kingdom; Academic Unit of Gastroenterology, Royal Hallamshire Hospital, Sheffield, United Kingdom

**Author notes:** Corresponding author: Rachel Hawkins, BSc (Hons), MSc, The University of Sheffield, Sheffield Centre for Health and Related Research, School of Medicine and Population Health, 07814452433. **Financial disclosures:** Funding from The University of Sheffield PhD Research Scholarships is acknowledged in supporting RLH to conduct this review. **Conflicts of interest:** AJL has acted as a speaker or consultant for Takeda, BMS, Sandoz, JNJ, Celltrion and Medtronic. **Data availability:** Data available in supplementary material.

**Keywords:** Inflammatory Bowel Disease, avoidable unplanned admissions, healthcare inequalities, scoping review

## Abstract

**Background:** Access to healthcare may be driving unplanned and potentially avoidable hospital admissions for people diagnosed with Inflammatory Bowel Disease (IBD). Interventions to reduce unplanned and potentially avoidable admissions need to be developed based on a clear conceptual framework that identifies the system-level access barriers contributing to these admissions. This scoping review aimed to synthesise the health system components for reducing unplanned IBD admissions to develop a conceptual framework to guide future interventions for reducing unplanned admissions.

**Methods:** A scoping review was conducted to identify literature exploring factors associated with unplanned IBD admissions and interventions to reduce IBD admissions. Literature published between January 2000 and October 2024 was identified from four electronic databases (Medline, Embase, CINAHL and Pubmed). A narrative synthesis presented the findings, guided by Candidacy Framework, to understand issues in healthcare access.

**Results and conclusions:** Of 1980 records identified, 17 were included. Avoidable IBD admissions result from inequity across the patient journey through healthcare specifically in access to: (1) earlier intervention during a flare, (2) specialist clinical advice about symptoms and psychosocial issues, (3) rapid access to outpatient care, (4) patient education, (5) systems that support self-management, (6) proactive care strategies, and (7) collaborative health professional working and referrals. Addressing service permeability (ease of using services) and local production of candidacy (patient-provider relationships and macro-structural conditions) are understood as most important for addressing avoidable unplanned IBD admissions. The Health System Access Framework is useful for understanding how services need to address patient care.

**Lay summary:** Many unplanned hospital admissions for inflammatory bowel disease may be avoided. This review found that quicker access to specialist advice, proactive care, and patient education is crucial. This study developed a framework to help healthcare providers improve services and reduce these admissions for patients.

## Introduction

Inflammatory Bowel Diseases (IBD) are complex and relapsing conditions of the gastrointestinal tract (1). Many people living with IBD (PLwIBD) endure repeated hospital encounters (1,2). In the UK, 1 in 7 PLwIBD are diagnosed during emergency hospitalisation (3) with rising rates of unplanned hospitalisation for PLwIBD (4–6). Unplanned IBD admissions have profound social and psychological impact (7) and should be considered a healthcare priority (8). Unplanned IBD admissions account for high healthcare costs that are comparable to major diseases such as cancer and heart disease (9–11). Understanding how some admissions for IBD may be avoided is therefore important.

The definitions of avoidable and preventable admissions in chronic conditions are varied and poorly defined (12–14) which is problematic when attempting to address avoidable IBD admissions. Improper definitions of concepts is a common issue in healthcare research whereby healthcare concepts often have similar and overlapping meaning (15,16). For example, inconsistencies in definitions of ‘emergency’ and ‘urgent’ abdominal surgery for IBD have been previously highlighted (17). It is unclear if there are distinct differences in how terms such as ‘avoidable’ or ‘preventable’ IBD admissions are being used.

Clarifying terms, and the components of avoidable admissions in IBD through a conceptual framework could support future intervention development, healthcare providers decision making, more effective distribution resources, and importantly more clearly and defined future research tackling this issue (13,18).

Patient access to healthcare is a common theme across all avoidable admissions (19,20) and a prominent issue for PLwIBD (21). The Candidacy Framework (CF) (22) is an established framework for understanding inequalities in healthcare access (23,24) and in the emergency care setting (25). Across seven interrelated constructs (outlined in Table 1), the CF describes the patient journey through healthcare and eligibility for care incorporating individual, interpersonal, organisational and structural contexts of care access (Dixon-Woods et al; Liberati et al., 2022). Candidacy has been applied in understanding access to cancer services (24), emergency care (25) and more recently in rheumatoid arthritis care (26)). Candidacy describes how a person must first recognise a health need (identification), know how and where to seek help (navigation), gain entry to services (permeability), present their case to a clinician (appearance), and receive a professional judgment about eligibility for care (adjudication). Patients may then accept or resist what is offered, and all of these stages are shaped by local operating conditions such as workforce capacity, service policies and existing patient-provider relationships. It is the cumulative effect of barriers across these stages, rather than any single factor, that the Candidacy Framework captures.

**Table 1.** Stages of Candidacy adapted for inflammatory bowel disease healthcare.

| **Candidacy Construct** | **General description from Dixon-Woods et al. (2006)** | **Adapted description to IBD healthcare** |
| --- | --- | --- |
| **Identification of Candidacy** | How people come to recognise symptoms as needing medical intervention. | PLwIBD identifies themselves as a candidate for IBD care. This is influenced by their social contexts, prior experience and knowledge of bowel symptoms, IBD and healthcare. |
| **Navigation of IBD services** | To access services, people must be aware of them and have available practical resources to access them. | Requires awareness of care options, mobilisation of resources (e.g. transport to clinics) and understanding of how the service manages IBD. For example, knowing that IBD flare advice lines are a first point of call during a flare. |
| **Permeability of IBD services** | Refers to the ease at which people can use health services, which is impacted by the organisation of services. This may include accessing appointments or referrals. | Represents PLwIBD journey through the system. The permeability of IBD services, i.e. the availability of and barriers to expert advice in primary or secondary care. |
| **Appearance at services** | When appearing at services people must have the competence to articulate their issue and assert their claim for medical intervention. | PLwIBD must have the skills and confidence to present themselves to clinicians. This requires competencies in articulating IBD needs, and an understanding of IBD treatment options. |
| **Adjudications by healthcare professionals** | Professional judgments are made by the healthcare professional about candidacy and their eligibility for care. This then strongly influences subsequent access to healthcare and interventions. | Candidacy also requires judgement and decision making of IBD health professionals. The clinician identifies the patient as a candidate for interventions. Requires assessment of disease status and severity of IBD flares. |
| **Offers of, and resistance to services** | Some people may choose to refuse offers or there may be resistance to healthcare and interventions such as referrals and medications. | This stage involves decision-making of PLwIBD to accept new treatments (e.g. biologics, steroids, surgery) and attend appointments. This is influenced by individual, social and organisational factors including trust in health care professionals and institutions. |
| **Operating conditions and local production of candidacy** | These include factors at the societal and macro levels that influence candidacy. For example, the availability of local resources and the relational aspects between the healthcare provider and the patient. | Represents the impact prior and current relationships PLwIBD have with their IBD team which impact Candidacy. Access to IBD care is also impacted by the availability of specialist IBD resources, and local and national policies that impact funding of services. |
| *PLwIBD = People living with inflammatory bowel disease* |  |  |

Prognostic reviews predicting admissions have shown younger onset, disease location and type of disease (e.g. stricturing), inadequate pain control and previous unplanned surgery predict IBD admissions (5,27). Concomitant psychological conditions also increase the likelihood of IBD hospitalisations (28–30). The additional contribution of ethnicity, deprivation and rurality suggest that an intersectional health inequalities approach is necessary (21).

IBD healthcare services encompass complex interrelated systems, including specialist, multidisciplinary care, primary care and community care (3,31). These complex health systems include multiple factors, organisations, processes, and people that influence care across the patient journey (21,32). However, very little is known about *what* and *how* components of the IBD healthcare system interact and may impact IBD admissions that could be prevented. Avoidable IBD admissions may result from failures elsewhere within the system, but there is no formal conceptualisation of this. It may be difficult to define an avoidable IBD admission using a single parameter (e.g. prognostic or psychological alone (32), due to the complexity of factors that may lead to a patient being admitted (33,34). The development of a conceptual framework of avoidable IBD admissions may therefore provide a structure to inform service improvement interventions on preventing these types of unplanned IBD admissions.

This scoping review aimed to provide an overview of the current research and address these gaps by developing a comprehensive conceptual framework of avoidable IBD admissions from a systems perspective. To do this, descriptions of concepts, factors and interventions associated with avoidable unplanned IBD admissions were synthesised. Scoping reviews are appropriate when examining a broad and complex topic and when the existing knowledge is heterogenous and yet to be comprehensively reviewed (35,36).

The Candidacy Framework (CF) (22) was chosen to guide this work due to its established utility in understanding inequalities in healthcare access across various conditions and contexts, including emergency care settings. The Candidacy Framework is primarily a model of healthcare access rather than a model of health inequity. It conceptualises the stages through which individuals negotiate access to healthcare services, identifying how organisational, interpersonal and structural factors create barriers at each stage (22). These barriers to access may disproportionately affect certain populations, thereby producing or reinforcing health inequities. In this review, the Candidacy Framework was used to identify the structural and system-level causes of barriers to IBD care that may lead to avoidable admissions. This builds directly on a prior scoping review by the authors (21), which identified broad inequalities in IBD healthcare, including higher rates of avoidable admissions among vulnerable groups. This review did not specify the health system failures contributing to these disparities. The present review moves from describing inequalities to analysing their structural causes within IBD care systems.

This review aimed to develop a conceptual framework of avoidable IBD admissions. To achieve our aim of developing a conceptual framework, we addressed three interconnected research questions (RQ):

1: What are the healthcare components used in definitions and descriptions of associated terms of avoidable unplanned admissions?
2: What health system factors and interventions have been evidenced for avoiding unplanned IBD admissions?
3: How can the Candidacy Framework help understand issues with access in IBD healthcare related to unplanned IBD admissions?

While a prior scoping review by the authors identified broad inequalities in IBD healthcare, including a higher rate of avoidable admissions in vulnerable groups (21), it did not specify the health system failures contributing to this issue. This review therefore moves from identifying the problem to analysing its causes from a systems perspective. We aim to develop a targeted framework of the specific healthcare components and interventions that can prevent avoidable IBD admissions.

## Materials and Methods

The full protocol for this scoping review is registered on Open Science Framework (https://osf.io/u5jm4/) and is reported according to PRISMA for scoping reviews (PRISMA-ScR).

### Eligibility criteria

Eligibility for study inclusion followed the population, concept and context (PCC) criteria, which stated:

⍰ **Population:** Adults (>16 years) with a confirmed IBD diagnosis, including with co-morbid physical and/or mental health conditions.

⍰ Exclusion: Pregnant IBD populations, due to unique healthcare needs of this IBD subpopulation.
⍰ **Concept**: Factors or interventions for unplanned IBD admissions, readmissions, and/or emergency room visits described as avoidable, preventable, modifiable or similar.

⍰ Exclusion: Basic prognostic factor studies, vaccination prevention infections resulting in admissions.
⍰ **Context**: Health system factors within outpatient or community care associated with unplanned hospital admissions, emergency department visits and readmissions (including related terms). The focus on health system factors rather than individual patient predictors (excluded) reflected the study’s aim to develop a conceptual framework that can inform service improvement interventions at the system level.

⍰ Exclusion: Preventing in-patient hospital outcomes, reporting on healthcare utilisation, healthcare expenditure, unnecessary examinations, surgeries and therapies.
⍰ Exclusion: Reducing or avoiding outpatient visits, GP visits, without specifying unplanned admissions.

Papers were required to be fully accessible in English and published from the years 2000 to 2024. Papers published prior to 2000 were excluded due to these studies reflecting a pre biologic era for IBD therapy (37). Peer-reviewed primary research articles were included. Excluded articles were basic science, case reports, conference abstracts, commentaries, and studies focused on individual patient predictors of admissions (5,27).

### Search strategy

A search strategy (insert link to supplementary file A), combined MeSH and free-text terms related to IBD (“exp crohns disease/”, “exp ulcerative colitis/”) and for terms related to avoidable admissions (“avoidable”, “preventable”, “unplanned”). The strategy was iteratively refined as new papers were identified through pearl growing and reference list screening.

### Information sources

First, MEDLINE, Embase via Ovid, CINAHL via EBSCO were searched following the search strategy. Then, additional Google Scholar searches were run to identify additional grey literature. Reference lists from relevant systematic reviews were also screened, along with targeted PubMed search to check for missed articles (38).

### Selection of sources for evidence

The initial search was conducted by [initials removed], and [initials removed] conducted a second search acting as a double reviewer. Search results were exported to Rayyan.ai (https://www.rayyan.ai/) for duplicate removal, title and abstract screening. Articles identified to be relevant to RQs 1-3 were then full text reviewed. If eligibility was unclear following full-text screening, for example where it was unclear if articles explicitly referred to the concept groups of interest (synonymous terms for avoidable admissions), these were excluded. Decisions regarding selection of unclear articles were made during project discussion meetings between [initials removed].

### Data charting and items

A matrix for chartering data extracted from the articles were created based on the following items:

1. Article characteristics: Tabulated study author(s), year of publication, country, standardised study design (39), research setting, study population, and total sample size.
2. Concepts of IBD admissions: Key terms used in articles (e.g., preventable, avoidable, unplanned, unnecessary) and any definitions.
3. Factors and interventions reported: Summary of findings, factors associated with admissions, interventions (if applicable), and the relationship (positive, negative, no association) with IBD admissions.
4. Candidacy Framework analysis: Deductive coding of extracts from included articles.

Data charting was conducted by one reviewer [initials removed], checked by a second [initials removed] and discussed with the wider team at team meetings [initials removed]. A critical appraisal of the included articles was not conducted as critical appraisal is not required in scoping reviews. The main function of scoping reviews is to describe the body of literature (40).

### Approach to analysis

A qualitative synthesis was conducted to better understand issues of context and complexity, offer insights that could support theory generation and inform clinical practice (41). The process involved: 1) tabulating evidence; 2) thematic synthesis; 3) development of a context-specific model within the CF; and, 4) identification of research gaps and future research priorities. The final framework aimed to illustrate both the healthcare components that define avoidable IBD admissions and how these components interact with patients’ journeys through the healthcare system using the CF.

*RQ1:* Healthcare components in descriptions of related terms of avoidable unplanned IBD admissions

A thematic synthesis of the descriptions of IBD admission concepts was first conducted (42), and used to build the initial framework, which is visually represented in the final model (Figure 2). First, it was explored if different concepts (e.g. reducing, preventing, avoiding) had the same or distinct meaning (16). This was performed to determine if it mattered which terms were used in the conceptual framework. Then, themes capturing the descriptions were captured in a “Patterning Chart”, as used in other conceptual scoping reviews (43). The Patterning Chart used a systematic process of coding and categorisation of all descriptions of avoidable (and related terms) unplanned IBD admissions. For each article, healthcare components were tabulated and counted, creating a matrix that visualised patterns of their descriptions.

This analysis of terminology was not merely as a semantic exercise but a necessary foundation for developing an actionable framework. By understanding how different admission terms are used in the literature we can ensure that our conceptual framework encompasses all relevant aspects of care that might influence unplanned admissions. The patterning chart described the conceptual characteristics of the descriptions of unplanned IBD admissions. A qualitative synthesis accompanied the Patterning Chart which describes concepts in a working framework of avoidable IBD admissions.

*RQ2: Synthesis of factors and interventions evidenced in avoiding unplanned IBD admissions*

Factors and interventions evidenced in articles that help prevent unplanned IBD admissions were synthesised using two approaches. First, findings were categorised inductively from the literature. Using this data, explanations were developed of the relationships between different factors. These insights were integrated into the concept framework from RQ1.

*RQ3: Applying the Candidacy Framework to understand access to IBD care and admissions*

To develop a context-specific model within the CF, a conceptual framework was iteratively developed that mapped healthcare components of avoidable IBD admissions to the CF. To explore how patients access IBD healthcare, refined statements were used (refined and reviewed by [initials removed]) about IBD care mapped to the CF (Table 1). This has been previously applied in other published research (24). Articles were exported to NVivo and codes relating to accessing care and IBD admissions were generated. Key themes from the literature regarding avoidable admissions were organised according to the stages of candidacy (identification, navigation, permeability, etc.). Findings were mapped to seven key statements about IBD care and to the conceptual framework of avoidable IBD admissions. This analysis helped refine the working framework from RQ1 and RQ2 into a conceptual framework.

## Results

### Study selection

Searches retrieved 1980 records, from which 257 duplicates were removed, 1566 records were excluded based on their titles, 81 based on their abstracts, and 59 based on their full-text (Figure 1). The remaining 17 articles were included in the review.

**Figure 1.**
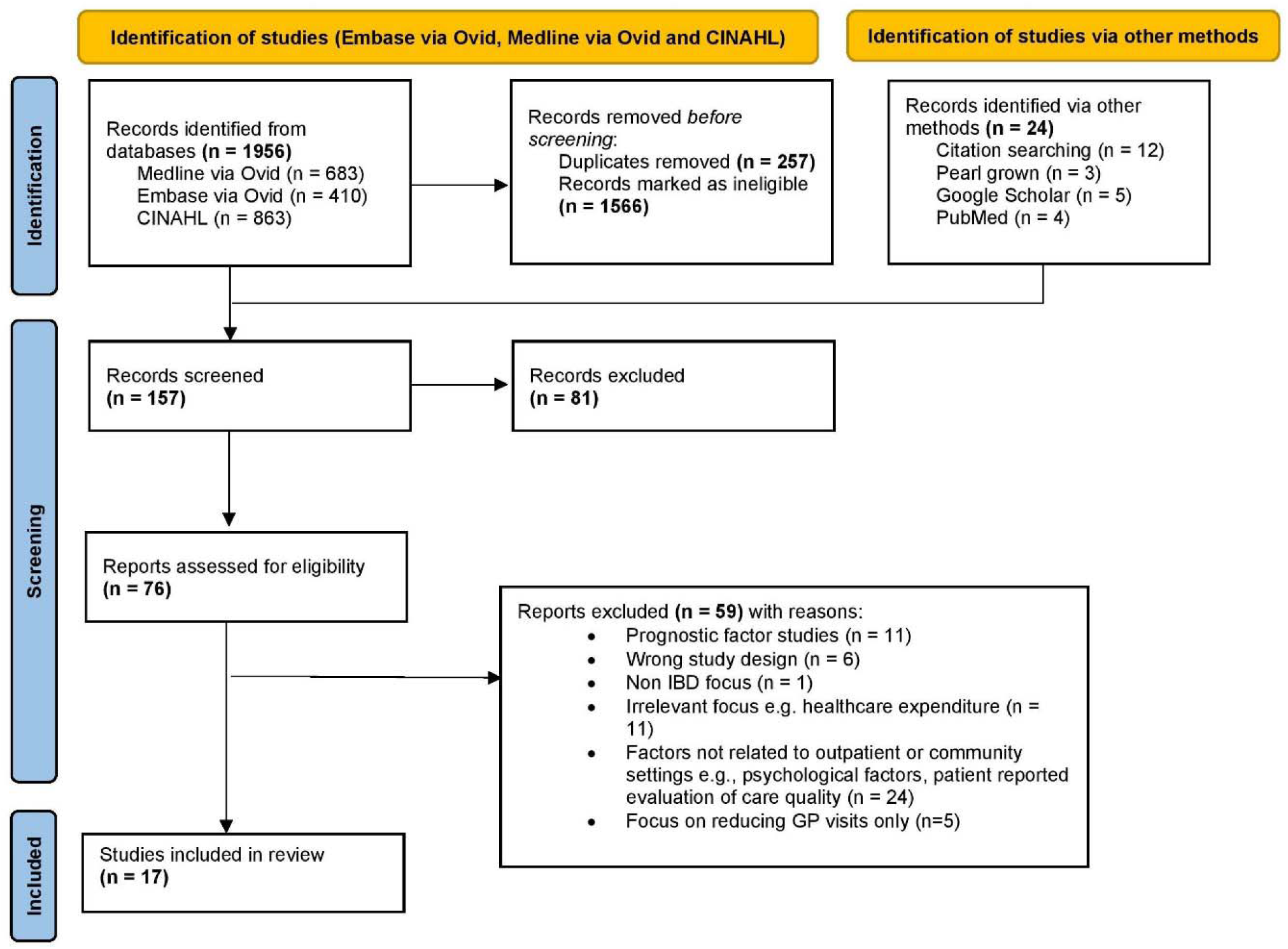
PRISMA flow diagram.

### Characteristics of included articles

Included studies (outlined in Table 2) were from: The USA (n=5) (44–48); Australia (n=3) (49–51); Canada (n=3) (52–54); the UK (n=2) (8,55); Belgium (n=1) (56); the Netherlands (n=1) (57); Israel (n=1) (58); and France (n=1) (59). The study designs of included studies were: Retrospective cohort studies (n=10) (45–51,54,58,59); prospective cohort studies (n=2) (44,56); cohort studies (n=2) (52,53); a survey (n=1) (55); a randomised controlled trial (n=1) (57); whilst one record was a UK charity report (8). Thirteen studies involved people with CD, 15 with UC and one with IBD Unclassified (55). Three studies did not report the IBD subtypes (8,47,50). Where reported, sample sizes ranged from 32 to 20,382. Three of the seventeen included articles reported the ethnicity of their samples (45,46,48), showing a proportionally higher White/Cucasian samples compared to other ethnic groups reported. The majority of articles also did not report socioeconomic characteristics of study samples (e.g. deprivation, income, education). Educational level (57) and English proficiency (48) was however reported by two studies.

**Table 2.** Characteristics and summary of included articles.

| Author(s),<br>year, country<br>of study | Study design | Setting | IBD population | Sample size | Outcome; Intervention detail or factors<br>investigated and summary of findings |
| --- | --- | --- | --- | --- | --- |
| Melmed et al.<br>(2021), USA | Prospective<br>cohort | GP, academic centres &<br>private community<br>practices | 54% female, 46% male<br>CD (58%), UC (38%), “other” (4%)<br>Ethnicity: Does not report<br>Socioeconomic (SES)<br>characteristics: Does not report | 20,382 | Reduced admissions: Implemented a toolkit of<br>19 service change interventions tied to<br>primary and secondary care e.g. patient<br>education materials about how to seek urgent<br>care, reserved clinic slots, rapid return calls,<br>rapid access clinics. |
| Nene et al.<br>(2020),<br>Canada | Observational<br>cohort | Hospital: tertiary IBD<br>centre | 41.3 % male, 58.7% female<br>CD (68.4%), UC (31.6%)<br>Ethnicity: Does not report<br>SES characteristics: Does not<br>report | 488 | Reduced admissions: Through a rapid access<br>clinic (RAC) with an IBD nurse or clinician for<br>patients with urgent clinical needs. The rapid<br>access clinic served as a mechanism for fast<br>tracking evaluations and treatment decision<br>making, when compared to accessing support<br>via the ED care. |
| Regueiro et<br>al. (2018),<br>USA | Retrospective<br>cohort | Hospital: University of<br>Pittsburgh | 57.3% female, 42.4% male<br>CD (62%), UC (38%) | 322 | Reduced admissions: A series of service<br>changes which is composed of: 1) stratifying<br>determinants of healthcare use, 2) |
|  |  |  | Ethnicity: 81% Caucasian, 2.5% Black, 2% Asian, 2% Middle Eastern, 1% Indian, 14.9% unknown<br>SES characteristics: Does not report |  | implementing the use of care plans and 3) a collective care health plan. The programme was led by the gastroenterologist and psychologists, supported by a social worker, dietitian, nurse coordinators, appointment coordinators and advanced practice providers. |
| Malhotra et al. (2018), USA | Retrospective cohort | Hospital: Minneapolis | 96.2% male<br>CD (56.9%), UC (43.1%)<br>Ethnicity: Caucasian (90.8%)<br>SES characteristics: Does not report | 130 | Increased admissions: Patients without a follow-up visit with primary care or a Gastroenterologist was associated with greater unplanned readmissions |
| Coenen et al. (2017), Belgium | Prospective cohort | Tertiary IBD centre | 58% female, 42% male<br>CD (72%), UC (28%)<br>Ethnicity: Does not report<br>SES characteristics: Does not report | 1313 patient contacts | Reduced admissions: The introduction of an IBD nurse providing face-to-face contact, a telephone line and email service. The service supported quick access to advice and care. |
| Zhen et al. (2021), Canada | Observational cohort | Hospital: Gastroenterology clinic | 62.5% male, 37.5% female<br>CD (71.8%), UC (28.2%)<br>Ethnicity: Does not report<br>SES characteristics: Does not report | 32 | Reduced admissions: A Digital app intervention (HealthPROMISE) provided patients with access to a self-management intervention. Scores on the app enabled earlier intervention of deteriorating patients through prompt triaging of patients that needed urgent care. |
| Liu et al.<br>(2019), USA | Retrospective cohort | Hospital: Tertiary veterans health centre | Does not report for full sample, admission patients only<br>Ethnicity: Does not report<br>SES characteristics: Does not report | 604 | Reduced admissions: A service redesign which adopted a model consisting of a preventative health check, psychological care, nutritional support and pain management. |
| de Jong et al.<br>(2017),<br>Netherlands | Randomised controlled trial | Hospital: 2 non-academic hospitals | 58% female, 42% male<br>CD (60.6%), UC (39.4%)<br>Ethnicity: Does not report<br>SES characteristics: Educational level reported | 465 | Reduced admissions: A digital intervention (myIBDcoach) included collecting PROMS 1) self-monitoring of symptoms 2) a personalised care plan 3) e-learning patient education modules and 4) communication with IBD clinic via the admin office |
| Harvey et al.<br>(2021),<br>Canada | Retrospective cohort | Hospital: secondary care centre | 60% male, 40% female<br>UC (78.5%), Pancolitis (21.5%)<br>Ethnicity: Does not report<br>SES characteristics: Does not report | 140 | Reduced admissions: A service intervention which added early evaluation delivered by a specialist IBD nurse following 2 weeks of new course of corticosteroid therapy. Early evaluation included education about the drug and nurse helpline information. |
| Leach et al. | Retrospective | Tertiary IBD centre | 55.4% female, 54.6% male | 556 | Reduced admissions: Service change |
| (2014),<br>Australia | cohort |  | CD (61%), UC (25.3%), non-IBD (13.7%)<br>Ethnicity: Does not report<br>SES characteristics: Does not report |  | intervention which implemented a nurse-led telephone and email intervention |
| Gethins<br>(2020), UK | Survey | Hospital: IBD service | Does not report gender<br>UC (60%), CD (38%), Unclassified (2%)<br>Ethnicity: Does not report<br>SES characteristics: Does not report | 50 | Increased admissions: Were as a result of IBD providers not initiating patients on a treatment linked to an acute exacerbation, as well as a lack of urgent face-to-face review. |
| Sack et al.<br>(2012),<br>Australia | Retrospective cohort | Hospital: IBD service | Does not report gender/subtype<br>Ethnicity: Does not report<br>SES characteristics: Does not report | 451 | Reduced admissions: A service intervention introducing a new formal IBD service consisting of: medical–surgical clinic, designated clinics, helplines, follow-up symptomatic clinics, patient education, regular radiology reviews and blood test monitoring |
| Martinez-Vinson et al.<br>(2020),<br>France | Retrospective cohort | Hospital: IBD service | 56% female, 44% male<br>CD (87%), UC (13%)<br>Ethnicity: Does not report<br>SES characteristics: Does not report | 252 | Reduced admissions: A service change which introduced an IBD nurse who provided help and advice to patients, and had a formal role in patient education. |
| Karimi et al (2021), Australia | Retrospective cohort | Hospital: IBD service | 52.6% female, 47.4% male<br>CD (66.5%), UC (30.5%)<br>Ethnicity: Does not report<br>SES characteristics: Does not report | 272 | Reduced admissions: A new service intervention which consisted of a IBD helpline, specialist nurses and virtual clinics |
| IBD UK 2021, UK | Charity report - survey | UK-wide charity report | Does not report gender/subtype<br>Ethnicity: Does not report<br>SES characteristics: Does not report | 10,222 | Increased admissions: As a result of patients not accessing specialist care and treatment quickly enough. Admissions occur as a result of missed opportunities for earlier treatment |
| Goren et al. (2022), Israel | Retrospective cohort | Tertiary IBD centre | 111 male (39.4%) and 171 female (60.6%)<br>188 (66.6%) with CD (33.3%), 94 with UC<br>Ethnicity: Does not report<br>SES characteristics: Does not report | 282 | Reduced ED revisits: A service change which consisted of an MDT IBD team (IBD dietitian, IBD nurse, psychologist, advanced endoscopist and colorectal surgery/proctology services) within the ED department which initiated specialised assessment referrals |
| Hassid et al. (2023), USA | Retrospective cohort | Non-profit healthcare organisation database | 914 (43.3%) male, 1197 (56.7%) female | 2111 | No effect on admissions found in patients who achieved a prompt outpatient follow-up after |
|  |  |  | 1065 (50.5%) UC, 1046 (49.6%)<br>CD<br>Ethnicity: Non-Hispanic White<br>1273 (60.3), Asian/Pacific Islander<br>205 (9.7), Black 172 (8.2),<br>Hispanic 326 (15.4),<br>Other/unknown 135 (6.4)<br>SES characteristics: English<br>proficiency (required interpreter)<br>10.8% recurrent ED visit group,<br>Neighborhood median household<br>income 20.3% recurrent ED visit<br>group. |  | an initial ED visit |

*RQ1:* Healthcare components in descriptions of related terms of avoidable unplanned IBD admissions

Articles used a range of concepts interchangeably when describing IBD admissions (Presented in supplementary file B, Table 1). The majority (n=12) referred to “reduce[ing]” types of IBD admissions, including reference to Emergency Department (ED) visits (44,51,52,56,58) or broader terms, such as “unplanned care” (44,45). Also used were terms of “avoiding”/ “avoidance”/ “avoidable”” admissions (n=9 articles), which were used at a greater frequency when compared to “preventing” / “preventable” (n=2 articles) admissions. Other terms used with the same meaning included “unplanned urgent care” (44), “without involvement of unplanned hospital attendance” (54) and reducing “need for inpatient care” (50).

Articles described the healthcare components of these admissions either through presenting propositions about the causes and potential mitigators of admission types (8,46,48–50,52,54,56), by theoretically interpreting outputs from their own study results (45,47,51,52,54,55,58,59), and/or by citing previous IBD literature (8,44,46,53,55–58). However, not all (7/18) articles defined concepts explicitly (8,46,49,50,53,56,58)). Overall, all these terms applied to unplanned IBD admissions (i.e. reduce, avoid, prevent) had overlapping meaning when descriptions were analysed. Therefore the components synthesised from all these terms were incorporated into the overarching framework.

Multiple potential factors for avoiding IBD admissions were identified and themed, as illustrated in the Patterning Chart (Table 3).

**Table 3.**
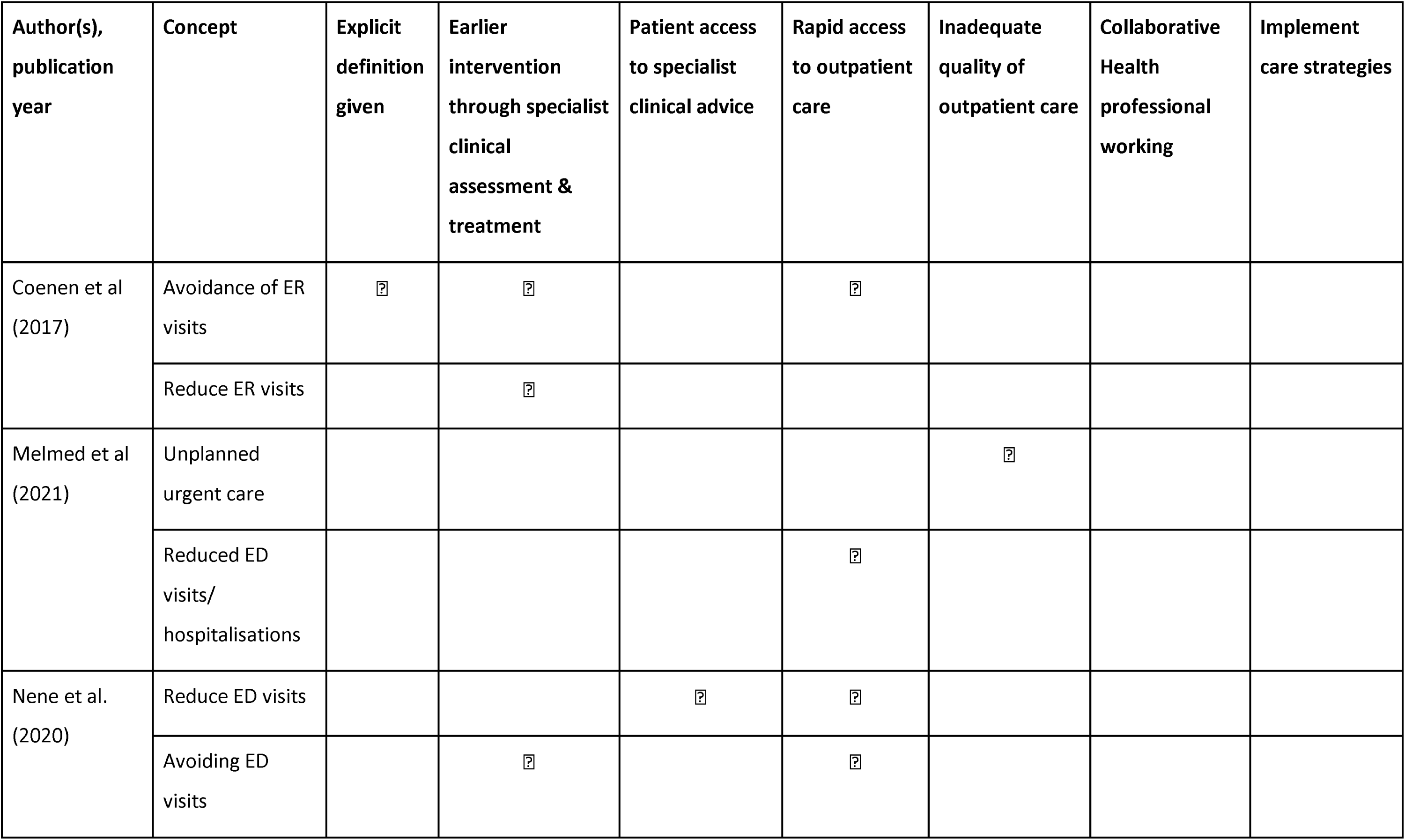
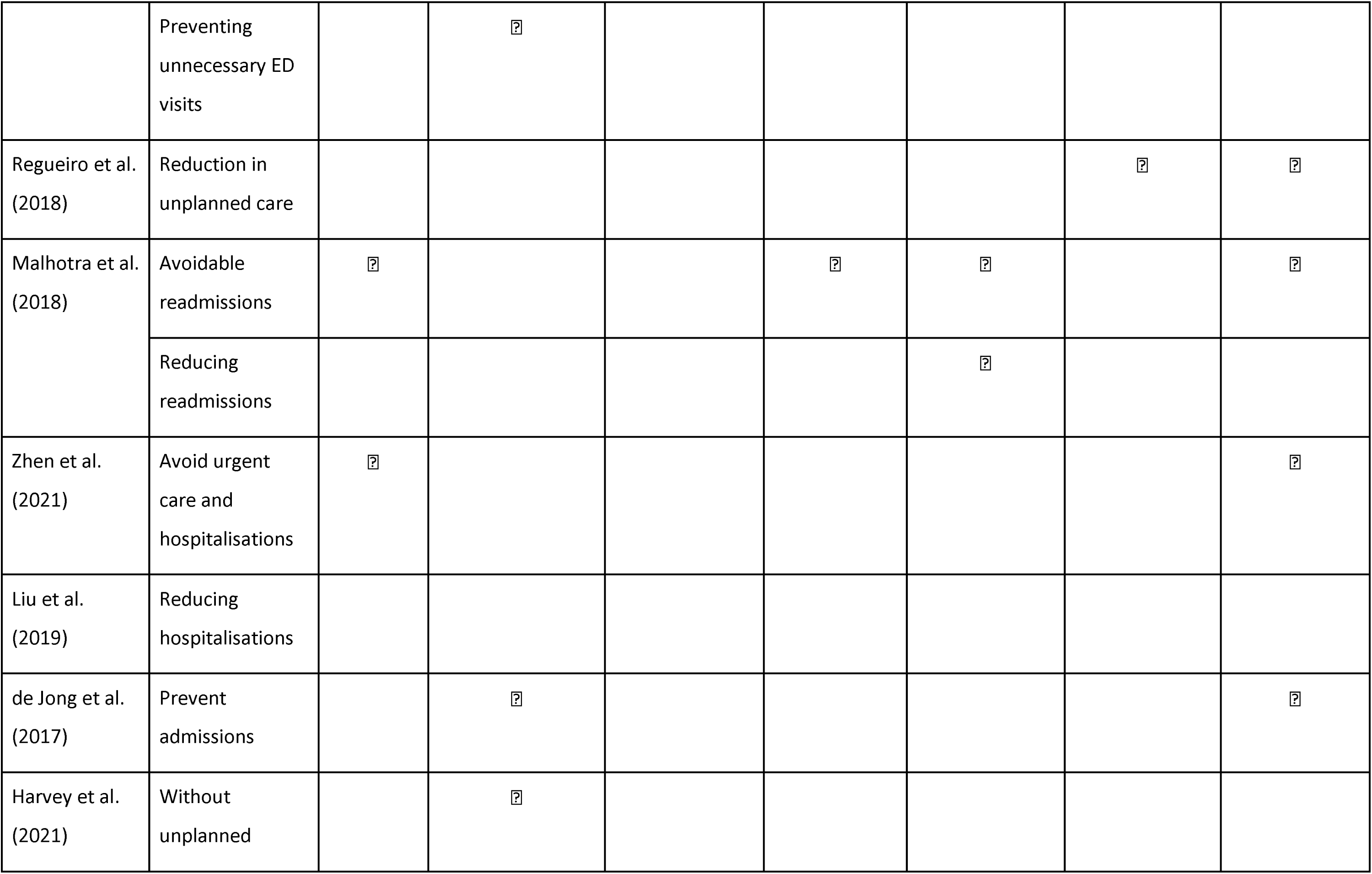
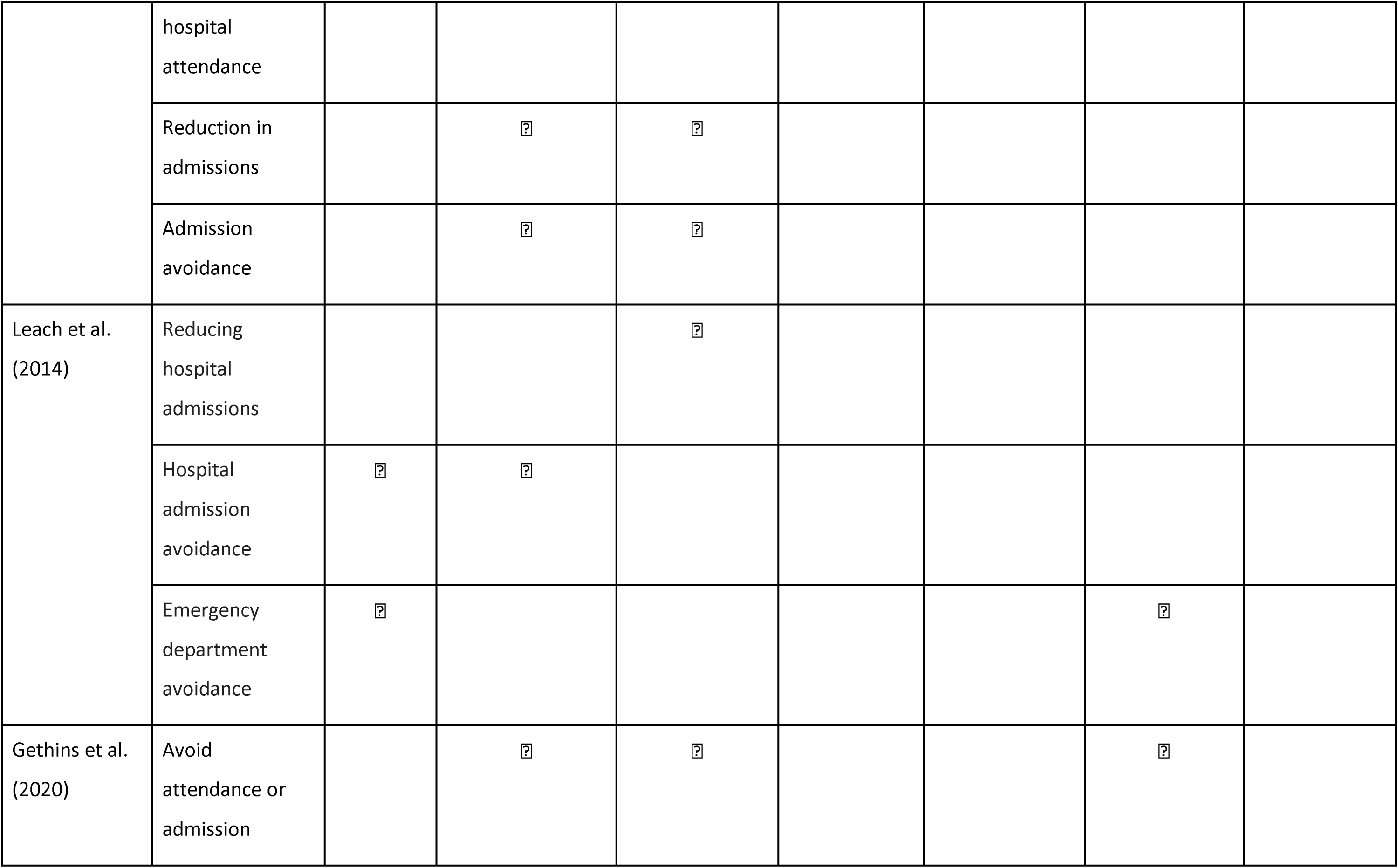
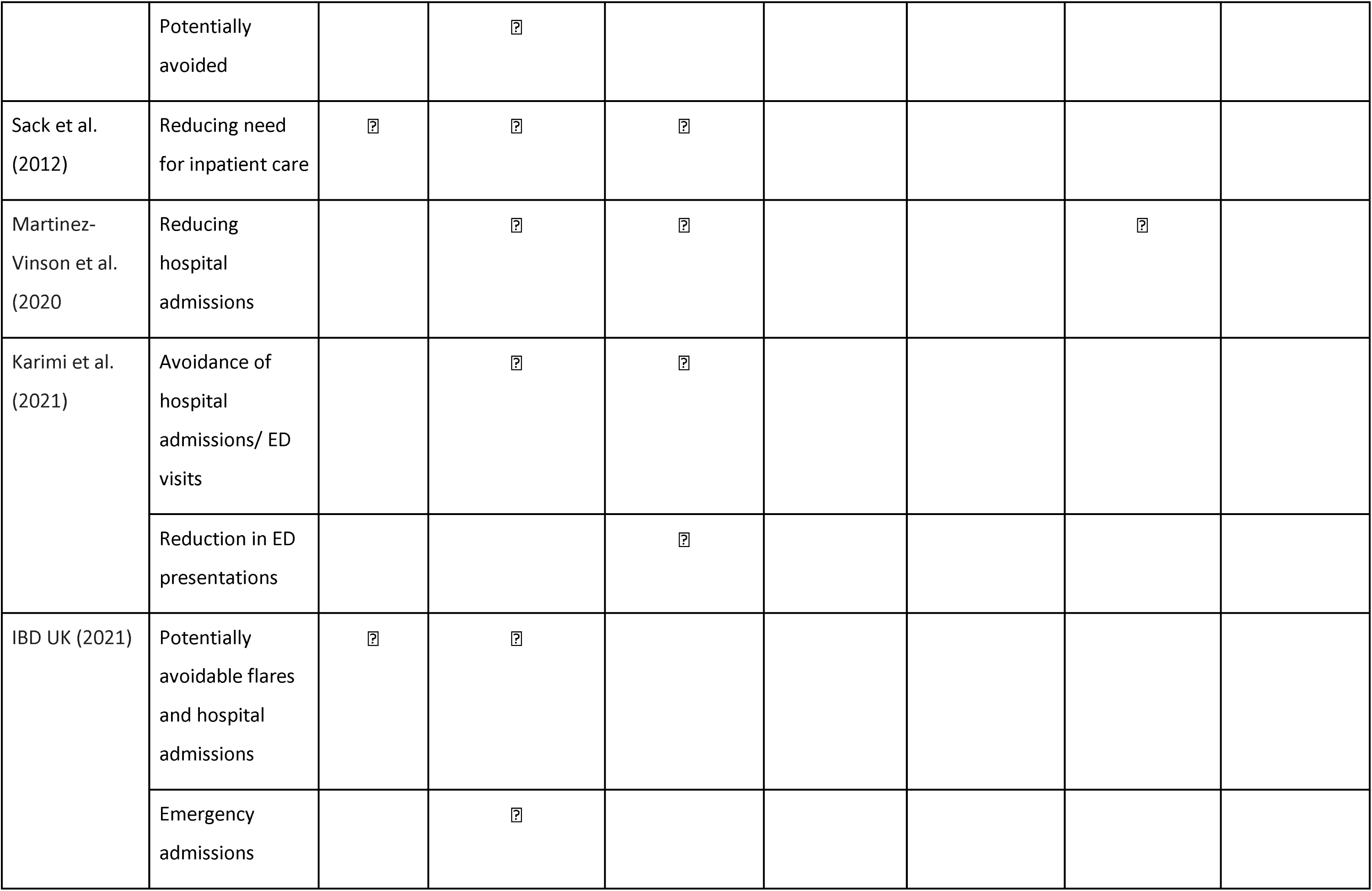
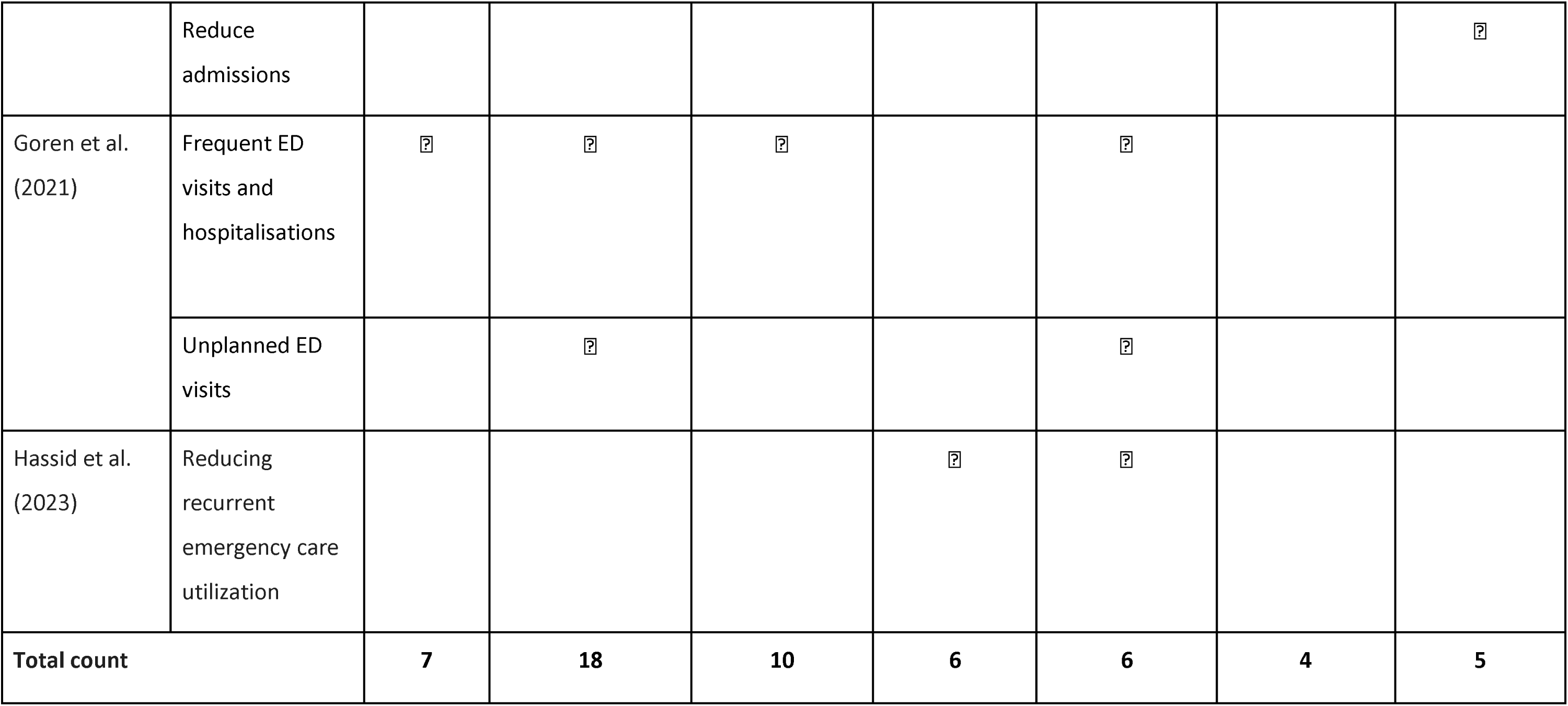
Patterning chart of the characteristics within theoretical statements describing IBD admissions.

**Table 4.** Outputs from included articles mapped to the Candidacy Framework.

| Candidacy construct | Inferences of access to IBD care and admissions<br>*Categories of improving patient access*<br>(study author, year) | Deductive mapping to conceptual framework |
| --- | --- | --- |
| Identification of candidacy | *IBD nurses and helplines*<br>IBD nurses are the first point of contact to facilitate prompt recognition of symptoms (49) and can empower patients to understand and recognise symptoms (56)<br>Use of an IBD helpline helps patients ask questions when in a flare, especially when newly diagnosed (Harvey et al., 2021) | Patient access to clinical advice |
|  | *Personalised care strategies*<br>Telemedicine tools support patient self-monitoring of symptoms (57) | Embedded systems supporting self-management |
| Navigation of healthcare services | *Education and self-management*<br>Education materials about when and how to seek urgent care (44) | Embedded patient education |
|  | *Outpatient care*<br>Use of an IBD helpline helps patients navigate the service when experiencing a flare (22,56) | Rapid access to outpatient care<br>Patient access to clinical advice |
| **Permeability of healthcare services** | *Outpatient care (organisation of clinics)*<br>More permeable services have: Reserved clinic slots, designated rapid access clinics (44,50)) and rapid return calls (44,52), early intervention services (54), nurse-led telephone and email clinics (49), providing rapid access to timely advice, diagnostic procedures, maintenance therapies and services.<br>*Outpatient care (delayed care)*<br>Less permeable services have: Long waiting times and non-specialist IBD assessments are existing barriers (52,55) which can disproportionately impact already underserved individuals who experience barriers in accessing care (53)<br>Lack of an appropriate outpatient follow up which impact later IBD admissions (46) | Earlier intervention through access to clinical assessment and treatment<br>Patient access to clinical advice<br>Rapid access to outpatient care |
|  | *IBD nurses and helplines*<br>IBD nurses promotes promote rapid access to treatments and relevant departments (8,55,56,59) and support patient access to advice and educational resources (51,59) | Earlier intervention through access to clinical assessment and treatment<br>Patient access to clinical advice<br>Embedded patient education |
|  | *Personalised care strategies*<br>Digital health monitoring platforms provide prompt access to urgent care (53) | Rapid access to outpatient care |
|  | and easy and accessible contact with IBD nurses (57) |  |
| **Appearance at services and asserting candidacy** | *Outpatient care (organisation of clinics)*<br>❑ RAC clinics supported patient appearance at services in comparison to appearing at ED departments (52) or if not face to face (55)<br>❑ An early intervention service supports patients to appear at services and assert their for healthcare (54) | Rapid access to outpatient care |
|  | *Personalised care strategies*<br>❑ Appearance at services is supported through communication with the clinic supported through the app (57) | Embedded systems supporting self-management |
| **Adjudications from healthcare professionals** | *Outpatient care (organisation of clinics)*<br>❑ RACs help healthcare professionals make optimised treatment decisions and to fast track evaluations (52), which are timely (55)<br>❑ Early intervention services help professionals to assess patients symptoms and response to steroids (22) | Earlier intervention through access to clinical assessment and treatment |
|  | *Personalised care strategies*<br>❑ Stratifying patients by biopsychosocial factors helps healthcare professionals allocate care plans and resources (45)<br>❑ Digital monitoring helps healthcare professionals identify deteriorating patients earlier (53) | Proactive care strategies<br>Earlier intervention through access to clinical assessment and treatment |
|  | *IBD nurses*<br>IBD nurses are key adjudicators of triage (56) which are sorted by clinical urgency, booking of routine clinic appointments and arrangements of diagnostic tests (51)<br>*Clinicians (non-specified)*<br>Management of IBD flares requires adjudications from professionals to evaluate the severity of illness and deliver a treatment plan (48) | Rapid access to care<br>Earlier intervention through access to clinical assessment and treatment |
| **Offers and resistance to services (patient decision making)** | *IBD nurses*<br>IBD nurses impact offers and resistance to treatments through education of risks/benefits and treatment adherence (56) | Patient access to clinical advice |
| **Operating conditions and local production of candidacy** | *Education and self-management*<br>Provision of patient education to support self-management of IBD (44,47,50,57) e.g. monthly patient education groups support patient empowerment (59) | Embedded patient education |
|  | *Patient-provider relationships*<br>Positive nurse-patient relationships support candidacy as nurses discuss personal and psychosocial impact (56), deliver patient-education, promote knowledge, discuss treatment goals and set patient expectations (47,49,52,55)<br>Continuous interaction helps with patient satisfaction with their relationships and interactions with providers (52) | Patient access to clinical advice |
|  | *Multidisciplinary care*<br>❑ Contribute to reducing IBD admissions in high healthcare utilisers (45) through preventative referrals and personalised healthcare (47)<br>❑ Psychologist and dieticians can support psychosocial coping of patients (59)<br>❑ Incorporating an MDT IBD team within the ED who conducts specialist assessment and interventions can reduce ED revisits, especially in service areas where there is generally poorer access to outpatient IBD care (58) | Collaborative health professional working and referrals |
|  | *Existing service organisation and policy*<br>❑ Presence of rapid return policies facilitate candidacy for access to care (44). Poor organisation of services and workforce constraints add to compounding pressures on outpatient clinic capacity (50,57)<br>❑ Services with higher patient numbers and lack of local resources may serve as barriers to implementing rapid access clinics/services (52) | Rapid access to outpatient care |

Six themes captured the components of the synonymous descriptions of avoidable IBD admissions, building the initial structure of the conceptual framework. Deficits in any of these six themed IBD healthcare components define an avoidable unplanned IBD admissions.

Avoidable IBD admissions result from *inadequate quality outpatient care (6 descriptions)*. The conceptual framework begins stating that reducing these admissions require health systems to:

1. **Intervene early to provide patients with access to specialist clinical assessment and treatment during a disease flare** (18 descriptions): Rapid access pathways enabled IBD specialists (Gastroenterologists, IBD nurses) to assess, evaluate, escalate and then intervene earlier which subsequently avoided an admission or ED visit.
2. **Provide timely patient access to specialist clinical advice about their symptoms or other psychosocial needs** (10 descriptions): IBD nurses and IBD helplines facilitate rapid access for PLwIBD seeking advice about their IBD symptoms or other psychosocial issues. PLwIBD seeking advice from IBD nurses support IBD specialists to assess, evaluate, escalate and intervene and avoid admission.
3. **Support rapid access to outpatient care** (6 descriptions): Regular access outpatient care encounters are necessary for high quality IBD care. Prompt outpatient follow ups can avoid hospitalisation and ED visits.
4. **Implement proactive care strategies** (5 descriptions): Groups several health system care strategies including use of proactive care, patient-centred approaches, care plans, risk stratified care and ED discharge planning.
5. **Embed multidisciplinary healthcare professionals to work together collaboratively** (4 descriptions): Collaborative and multidisciplinary working of IBD healthcare professionals is necessary for avoiding IBD admissions; this includes collaboration between tertiary outpatient care teams and ED teams.

*RQ2: Synthesis of factors and interventions evidenced in avoiding IBD admissions*

Most articles (13/17) reported on interventions that reduced IBD admissions by reshaping healthcare delivery. Interventions captured across the articles targeted quality improvement of services by improving patient access and experience (44,45,47,50,52,54,58), implementing a formal IBD nurse position (56,59), nurse-led helplines and virtual clinics (49,51) and digitally enabled patient monitoring interventions (53,57).

The remaining (4/17) reported on factors associated with IBD admissions; 2/4 studies focused on ED revisits and hospital readmissions (46,58). 3/5 evidenced factors that increased admissions (8,46,55), whilst one found no evidence of an association (58).

Interventions and factors investigated by studies concentrated on improving patient access to various elements of IBD care. Categories of improving patient access included to: outpatient care (44,50,52,57) including follow-up care (46,48), early intervention (8) and IBD nurses (49,51,54,56,59), prompt triage by providers (53–55), education and self-management (44,50,53,54,57,59) (new concept), multidisciplinary care (45,47,58), personalised care plans (45,57) and stratified care (45).

### Refinements to the Conceptual Framework

Two additional healthcare components were added to the conceptual framework (Figure 2). these healthcare concepts state that to reducing avoidable IBD admissions:

VI. **Require health systems to embed opportunities for patients to access education about the disease** (5 studies): Patient education programmes, patient empowerment for self-management, and education on how to access IBD services are necessary to equip patients with the knowledge to manage their condition and avoid admission.
VII. **Require health systems to embed systems that support patient self-management** (1 study): Digital tools facilitate self-management of patients and in this case can prompt recognition of need for urgent care.

**Figure 2.**
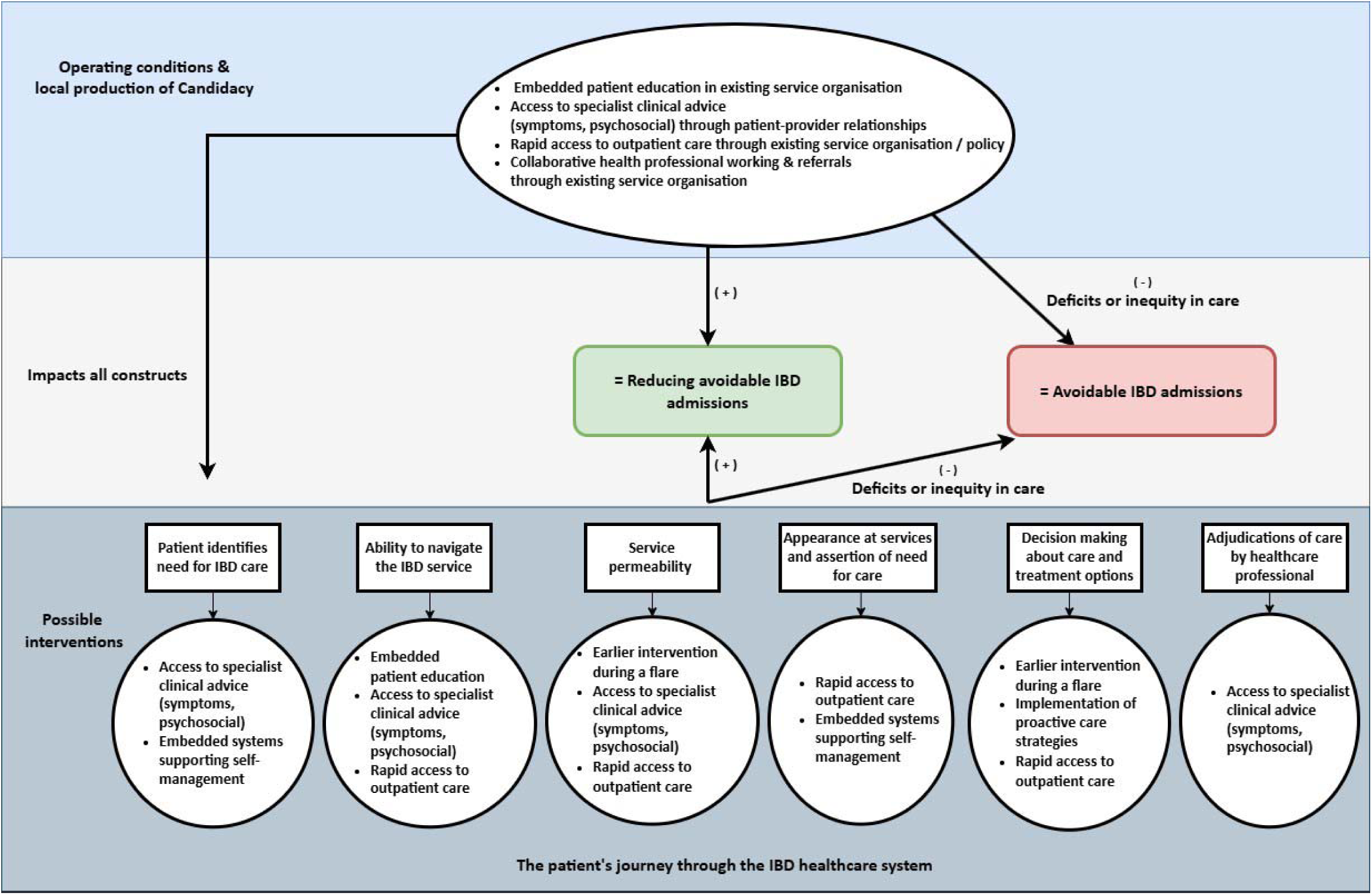
Framework presenting the healthcare concepts of avoidable IBD admissions. Themed concepts are mapped to the Candidacy Framework.

Additionally, data synthesised from the articles expanded the concept of “Embed multidisciplinary healthcare professionals to work together collaboratively”, which was updated to ‘Require multidisciplinary collaboration and referral systems’. This refined concept accounts for health professionals providing access to multidisciplinary referrals including psychosocial support services (psychological, pain management).

*RQ3: Applying Candidacy to understand access to IBD care and admissions*

Table 5 presents the tabulated inferences about IBD care and avoidable unplanned admissions that were extracted and interpreted using the Candidacy Framework and deductively mapped to the concepts of the conceptual framework developed from RQ1 and RQ2. Figure 2 presents the final Health System Access Framework for avoidable IBD admissions, integrating findings from all three research questions. Below provides an explanation of the conceptual framework of avoidable IBD admissions. Squared brackets at the end of paragraphs indicate the relevant concepts.

### Identification of Candidacy (mapped to 4 articles)

Before accessing IBD services, PLwIBD appraise their need for medical intervention (i.e. identify new symptoms). IBD nurses are key during this process who are the first point-of-call via IBD helplines (49,54) and empower PLwIBD to understand and legitimise themselves as candidates for healthcare (54,56).

Digital self-monitoring tools embedded within IBD services also facilitate patient self-management (57), supporting patients to identify candidacy [patient access to specialist clinical advice, embedded health systems that support patient self-management].

### Navigation of services (mapped to 3 articles)

PLwIBD must have the appropriate knowledge of their condition and of the IBD service in order to access healthcare. Patient educational materials support *Navigation of services* by informing patients about IBD and how to access the service during a flare (44). IBD nurses are key facilitators of navigation through delivering patient education and counselling, especially via IBD helplines (54,56) [health systems embed opportunities for patients to access education about the disease, patient access to specialist clinical advice, rapid access to outpatient care].

### Permeability of services (mapped to 13 articles)

PLwIBD journey through the healthcare system to reach the needed service and intervention. A more permeable IBD service reduces avoidable IBD admissions. This is achieved through better service organisation including reserved clinic slots, rapid access clinics and fast acting IBD flare helplines (8,44,50,52,54). Again, IBD nurses appear key in increasing service permeability, streamlining patients to access the appropriate intervention (49,51,54,56,59), as well as digital monitoring strategies (53,57).

Less permeable IBD services have long waiting times, lack of outpatient follow-up (8,46), and lack of access to specialist IBD assessments (55) [earlier intervention to clinical assessment and treatment, rapid access to outpatient care and patient access to clinical advice].

### Appearance at services (mapped to 4 articles)

Appearance and assertion of candidacy requires PLwIBD to articulate their needs to clinicians. Rapid access clinics support patients to assert candidacy (52,54), especially when compared to a patient appearing to a non-specialist in the ED (52). Telemedicine provides additional communication to IBD clinics (57) however barriers include lack of available face-to-face appointments (55) [rapid access to outpatient care, embedded systems that support self-management].

### Adjudications by healthcare professionals (mapped to 8 articles)

IBD health professionals decide if the patient is a candidate for accessing various forms of IBD care and these decisions are often timely. Nurses were notable decision makers for adjudications, triaging patients based on their assessment (48,51,54,56). Service organisation including rapid access clinics and digital monitoring support providers in making decisions about care through fast-tracking patients into the system (52,53,55). As well as services that utilise personalised care plans and early evaluation clinics to stratify decisions based on the patients needs (45,54) [earlier intervention to clinical assessment and treatment, require health systems to implement proactive care strategies, rapid access to outpatient care].

### Offers of, and resistance to services (mapped to 1 article)

PLwIBD must make a decision to accept treatment or attend appointments. IBD nurses appear key in supporting patients’ decision to take up treatments/appointments, playing a role in educating patients about the risks and benefits of treatment (56) [patient access to clinical advice].

### Operating conditions and local production of Candidacy (mapped to 12 articles)

Access to IBD care is shaped by the local health system operating conditions and existing patient-provider relationships. Within IBD care, Initiatives that fostered relationships (especially with nurses (47,49,55,56)) empowered patients were shown to support candidacy (52,54). The presence of local IBD service rapid return policies (44), rapid access clinics (44,55), multidisciplinary teams (45,58,59), and embedded patient education (44,47,50,57) are *local operating conditions* that support patient candidacy. Barriers include workforce constraints and compounding pressure on outpatient clinic capacity (50,52,57) [rapid access to outpatient care, collaborative health professional working, opportunities for patients to access education, patient access to clinical advice]

## Discussion

### Summary of findings and implications

This scoping review developed a conceptual framework of avoidable IBD admissions by synthesising the current evidence surrounding avoidable IBD admissions. The framework builds upon the six themes identified in RQ1, incorporates the additional healthcare components and refinements from RQ2, and organises these components according to the Candidacy Framework as analysed in RQ3. This integration illustrates how avoidable IBD admissions result from inequities in accessing healthcare across the patient journey and highlights the complex interplay between individual, interpersonal, and organisational aspects of IBD care by guidance from the Candidacy Framework (22). Each component of the framework represents a potential point for intervention to reduce avoidable admissions.

Findings from this review identified seven healthcare components - suggesting possible interventions. These included access to: (1) Earlier intervention during flares, (2) specialist clinical advice, (3) rapid access to outpatient care, (4) patient education, (5) self-management support, (6) proactive care strategies and (7) collaborative multidisciplinary care and referral systems (Figure 2). In practice, these components will overlap and are not distinct. We acknowledge that several of these individual components - such as timely specialist access, patient education and multidisciplinary care - are well-recognised principles of high-quality IBD care (31,60). The contribution of this review is not the identification of these components in isolation, but rather their synthesis into a unified, theoretically grounded framework that maps them to stages of the patient journey through healthcare. This integration provides a structured basis for designing and evaluating service-level interventions that address multiple components simultaneously.

New insights presented in this review can inform IBD health service commissioners and healthcare providers of possible interventions to reduce avoidable IBD admissions, adding new knowledge to the limited understanding (61). The findings argue that system driven avoidable IBD admissions results from deficits or inequity in the provision of IBD healthcare (see Figure 2), aligning with outputs from a previous IBD health inequalities review (21). We use the term ‘inequity’ here not to denote demonstrated disparities between specific demographic groups (which most included studies did not report) but rather to denote system-level deficits in access that have the potential to produce unjust differences in outcomes (62). Where services fail to provide timely specialist assessment, education or self-management support, patients who lack the resources, knowledge or relationships to navigate these barriers are most likely to experience avoidable admissions. The absence of sociodemographic reporting in most included studies is itself a limitation. Without such data, the extent to which these access failures differentially affect disadvantaged populations cannot be quantified from the current evidence base. Healthcare implementation research also recognises the structural factors of healthcare systems that impact unequal access, quality, or outcomes of care (63).

Upon exploring terms of “avoidable”, “preventable” and “reducing” IBD admissions, these were found to have similar and overlapping features, and were used interchangeably across articles (64). This lack of consistent terminology is also a recognised issue in research on Ambulatory Care Sensitive Conditions (ACSCs). (14,65). ACSCs are defined as “where effective community and person-centred care can prevent the need for hospital admission”(66). Moreover, recent scoping review research improving the understanding of the preventability of readmissions in heart failure also found significant heterogeneity in definitions and measures (67).

However, unlike ‘unplanned admissions,’ these terms align with the principles of values-based healthcare, which prioritises improving patient experience and outcomes through efficiently organised specialist care.. Values-based healthcare centres on equitable healthcare focused on improving patient experience and outcomes through efficient organisation of care with specialist expertise (68). Future research with those who make use of terms such as avoidable and preventable admissions, such as IBD clinicians, should explore the implications on research and patient care.

The framework developed in this study demonstrates that avoidable IBD admissions reflect inequalities in service access and suboptimal quality of outpatient care. IBD services must promote timely access to multidisciplinary care and referrals, to earlier intervention during flares, and to opportunities to empower and educate PLwIBD. The multi-pronged definition presented is similar to previous research defining avoidable ED presentations (69). Services must aim to address inequalities in; access to specialist assessments, specialist advice, outpatient care, and patient-centered care strategies. Thereby implying that IBD systems are failing to deliver IBD care according to policy standards (31).According to the evidence, IBD services that implement rapid access pathways, IBD nurse support, patient education, and self-management tools may influence admissions by enabling earlier intervention through improving patient knowledge and condition management. Consequently, IBD services and policymakers should consider these evidence-based components when designing interventions and allocating resources to IBD care.

Factors and interventions for reducing IBD admissions included improving access to; outpatient care, follow-up care, earlier intervention, IBD nurses, prompt triaging, education and self-management, multidisciplinary care, personalised care plans and risk stratified care. Inadequate access to timely and rapid IBD care appears to be associated with avoidable unplanned admissions. A potential mechanism of this, which should be explored, is that a lack of timely IBD care results in increased uncontrolled symptoms, escalating to an unplanned admission. PLwIBD often report inadequate experiences and access to IBD care (70–72). However, the appropriate timing and role of outpatient follow-up remains unclear, and warrants further investigation to understand its influence on reducing admissions (73). Future research is necessary to explore changes in the structure of care delivery which incorporate some or all of these mechanisms, to reduce unplanned care use (74).

Yet, despite improvements in patient outcomes reported by studies, persisting issues of patient access, experience, and outcomes of care continue (21). The current model of IBD healthcare is difficult to manage and maintain, impeding quality of care (3,75). In the UK, Crohn’s and Colitis UK (CCUK) have continuously called for faster access to referrals, specialist care and treatment (3,8). We recommend alternative approaches for improving IBD care which may be explored through qualitative research that aims to understand and prioritise new healthcare processes.

The application of the Candidacy Framework revealed that permeability of services and operating conditions and local production of candidacy were the most prominent constructs in the literature. This highlights that organisational factors—such as the ease of using services—and interpersonal aspects, like positive patient-professional relationships, are critically important in reducing avoidable admissions (76). The evidence repeatedly underscored the integral role of IBD nurses in supporting candidacy across the patient journey (49–51,56,59,60), yet most UK services do not meet recommended staffing levels, potentially creating a significant barrier to care (77) (78).

Conversely, less can be inferred about *offers and resistance to services and navigation of services,* which was anticipated given the review’s focus on system-level rather than individual factors like medication adherence (79,80). Intolerable side effects often make medication-use challenging (79). Future qualitative research is needed to explore these patient-centric aspects of candidacy. Furthermore, newer constructs, such as the embodied relational self from Candidacy 2.0, could offer deeper insights into how intersecting patient identities impact access and should be explored in future IBD research e (26).

No qualitative studies were found that identified these specific system-level factors and future qualitative research is recommended to better understand patient experiences.. Given the CF’s focus on inequalities in access to care within disadvantaged groups, future IBD research should also engage with underserved populations. It is well known that underserved groups which may be defined by ethnicity, race, gender identity, socioeconomic status, disability and other characteristics, experience significant inequalities in health (81). However, very few studies included in this review reporting on characteristics such as race/ethnicity, socioeconomic status or other underserved characteristics. This is an area of unmet need identified in another IBD review (21).

### Strengths and limitations

The application of Candidacy to IBD care is novel and a strength of this review. The CF supports an understanding of how patients negotiate access to healthcare services (22). Guided by the framework, we were able to systematically examine how various system components interact across the patient journey. This theoretical underpinning allowed us to move beyond isolated factors and towards more complex between healthcare access, interpersonal factors, service delivery, and avoidable admissions. Differences in health system orientation across countries should be considered when reviewing this evidence. All included studies originated from high-income countries with either universal public healthcare or insurance-based systems. Low-and middle-income countries (LMICs), where healthcare is frequently funded through out-of-pocket payments, were not represented. The concept of candidacy, particularly permeability and navigation, may operate very differently in settings where financial barriers determine initial access to any specialist care. Although the Candidacy Framework itself has been applied in LMIC contexts (82), the generalisability of our findings to LMIC contexts is uncertain, and future research should explore how out-of-pocket payment structures and resource-limited settings shape the patient journey to IBD care and potentially avoidable admissions.

Only two studies were eligible from the UK highlighting an evidence gap. The absence of primary care perspectives is also a limitation. Future research should explore the role of primary care providers, as interpersonal and organisational factors within these services are critical, especially given that significant diagnostic delays in IBD often originate at this stage of the patient journey(83).

Another limitation of this review is that the Health System Access Framework is only relevant to people with a confirmed diagnosis of IBD. Delayed diagnosis in IBD is a significant issue (83,84), which can result in emergency care outcomes including surgery (84). Recently published UK research showed that greater diagnostic delay resulted in more unplanned IBD admissions (85). Avoidable admissions in this population therefore requires further focused research to understand how delays to diagnosis result in potentially avoidable admissions. It is anticipated that additional health systems would be important to consider in this which are not represented in this review such as primary and community-based care services as illustrated in previous research of sources of diagnostic delay in IBD (83).

A key limitation is the absence of a formal quality appraisal, a standard practice for scoping reviews, which means the strength of evidence for each identified factor could not be formally assessed. Although absence of a quality appraisal is justified in scoping reviews (86), a mixture of methodologies and study designs were incorporated within this review. Furthermore, the high proportion of retrospective cohort studies means that while associations can be identified, causality cannot be inferred from this review. This warrants the need for future research to test these assumptions, to inform intervention design. These concepts should be viewed as potential explanatory variables within the system. Whether they are modifiable warrants future research across these areas to investigate at a systems level.

### Conclusion

This scoping review presents a conceptual framework outlining that avoidable IBD admissions reflect inequalities in service access and in suboptimal quality of outpatient IBD care. To address some unplanned IBD admissions, services must promote timely access to multidisciplinary care and referrals, to earlier intervention during flares, and to opportunities to empower and educate PLW IBD. Findings demonstrated that interventions to improve access to care and reduce interventions impact across the patient journey through healthcare, from identification of symptoms through to adjudications and treatment decisions made by healthcare professionals. Improving the ease of using healthcare services for PLW IBD, relationships with providers, and promoting access through the existing organisation of services are currently understood as most important for addressing unplanned IBD admissions. Further research is now necessary to explore and refine this access framework for IBD care.

## Data Availability

All data produced in the present work are contained in the manuscript

## Notes

### Summary of Updates

Term "inequity" has been updated to "access" in title of the framework. Further clarification on difference between this scoping rev iew and a health inequalities review we conducted in IBD care added.

